# Clinical and neurogenetic characterisation of autosomal recessive RBL2-associated progressive neurodevelopmental disorder

**DOI:** 10.1101/2024.05.03.24306631

**Authors:** Gabriel Aughey, Elisa Cali, Reza Maroofian, Maha S Zaki, Alistair T Pagnamenta, Fatima Rahman, Lara Menzies, Anum Shafique, Mohnish Suri, Emmanuel Roze, Mohammed Aguennouz, Zouiri Ghizlane, Saadia Maryam Saadi, Zafar Ali, Uzma Abdulllah, Huma Arshad Cheema, Muhammad Nadeem Anjum, Godelieve Morel, Robert McFarland, Umut Altunoglu, Verena Kraus, Moneef Shoukier, David Murphy, Kristina Flemming, Hilde Yttervik, Hajar Rhouda, Gaetan Lesca, Bibi Nazia Murtaza, Mujaddad Ur Rehman, SYNAPS Study Group, Genomics England Consortium, Go Hun Seo, Christian Beetz, Hülya Kayserili, Yamna Krioulie, Wendy K Chung, Sadaf Naz, Shazia Maqbool, Joseph Gleeson, Shahid Mahmood Baig, Stephanie Efthymiou, Jenny C Taylor, Mariasavina Severino, James EC Jepson, Henry Houlden

## Abstract

Retinoblastoma (RB) proteins are highly conserved transcriptional regulators that play important roles during development by regulating cell-cycle gene expression. RBL2 dysfunction has been linked to a severe neurodevelopmental disorder. However, to date, clinical features have only been described in six individuals carrying five biallelic predicted loss of function (pLOF) variants. To define the phenotypic effects of *RBL2* mutations in detail, we identified and clinically characterized a cohort of 28 patients from 18 families carrying LOF variants in *RBL2*, including fourteen new variants that substantially broaden the molecular spectrum. The clinical presentation of affected individuals is characterized by a range of neurological and developmental abnormalities. Global developmental delay and intellectual disability were uniformly observed, ranging from moderate to profound and involving lack of acquisition of key motor and speech milestones in most patients. Frequent features included postnatal microcephaly, infantile hypotonia, aggressive behaviour, stereotypic movements and non-specific dysmorphic features. Common neuroimaging features were cerebral atrophy, white matter volume loss, corpus callosum hypoplasia and cerebellar atrophy. In parallel, we used the fruit fly, *Drosophila melanogaster*, to investigate how disruption of the conserved RBL2 orthologueue Rbf impacts nervous system function and development. We found that *Drosophila Rbf* LOF mutants recapitulate several features of patients harboring *RBL2* variants, including alterations in the head and brain morphology reminiscent of microcephaly, and perturbed locomotor behaviour. Surprisingly, in addition to its known role in controlling tissue growth during development, we find that continued *Rbf* expression is also required in fully differentiated post-mitotic neurons for normal locomotion in *Drosophila*, and that adult-stage neuronal re-expression of *Rbf* is sufficient to rescue *Rbf* mutant locomotor defects. Taken together, this study provides a clinical and experimental basis to understand genotype-phenotype correlations in an *RBL2*-linked neurodevelopmental disorder and suggests that restoring *RBL2* expression through gene therapy approaches may ameliorate aspects of *RBL2* LOF patient symptoms.

## Introduction

Retinoblastoma (RB) proteins play well-defined roles in regulating cell-cycle gene expression during development^1^. The mammalian RB family consists of three members, RB1, RBL1 and RBL2, which share overlapping functions and have some exclusive roles as well. RB proteins antagonize the action of E2F transcription factors, which may result in the activation or repression of gene expression depending on the genomic context, and variants of RB proteins are linked to an array of disease states. For example, RB1 is a well-known tumor suppressor, with loss-of-function (LOF) mutations associated with several types of neoplastic lesions including retinoblastoma, prostate cancer, breast cancer, lung cancer and osteosarcoma^2–7^. RBL1 acts as a tumor suppressor by inhibiting the activity of E2F transcription factors, particularly E2F1, thereby preventing the inappropriate progression of cells through the cell cycle. RBL2 similarly functions as a key regulator of cell division through interactions with E2F4 and E2F5, and promotes senescence by repressing repair genes, controlling DNA methylation, and influencing telomere length^8–10^.

RBL1 and RBL2 have also been found to regulate neuronal differentiation and the survival of post-mitotic neurons^11^. Correspondingly, pathogenic variants in *RBL2* have been associated with severe developmental delay, dysmorphic features, microcephaly, and behavioural abnormalities^12–14^. However, clinical features associated with *RBL2* pathogenic variants have only been characterized in six individuals. Hence, a comprehensive characterization of this disorder has yet to be performed. Furthermore, the cell-types in which *RBL2* expression is required to promote neural development and function remain unclear.

To define the phenotypic effects of *RBL2* mutations in detail, we identified and clinically characterized a cohort of 28 patients from 18 families carrying homozygous or compound heterozygous predicted LOF (pLOF) *RBL2* variants. These studies have expanded the clinical spectrum and identified the most common dysmorphic and neuroradiological features linked to the disorder. Additionally, we have broadened the molecular spectrum by identifying fourteen new disease-causing variants, providing additional support for *RBL2* LOF as basis of this disorder.

*RBL2* null mice display embryonic lethality coupled with impaired neurogenesis and enhanced apoptosis^15^. Therefore, we used the fruit fly, *Drosophila melanogaster*, to investigate how disruption of the conserved RBL2 orthologueue Rbf impacts nervous system function and development. We found that *Drosophila Rbf* LOF mutants recapitulate several features of patients harboring *RBL2* variants, including alterations in the head and brain morphology reminiscent of microcephaly, and perturbed locomotor behaviour. Surprisingly, in addition to its known role in controlling tissue growth during development, we found that continued Rbf expression is also required in fully differentiated post-mitotic neurons for normal locomotion in *Drosophila*, and that adult-stage neuron-specific re-expression of Rbf is sufficient to rescue *Rbf* mutant locomotor defects.

Collectively, our work provides a clinical and experimental foundation to understand genotype-phenotype linkages in an *RBL2*-linked neurodevelopmental disorder, and suggests that restoring *RBL2* expression in post-mitotic neurons through gene therapy approaches may mitigate some of the morbidities caused by *RBL2* pLOF.

## Materials and methods

### Patient identification and genetic investigation

#### Patient recruitment

The affected individuals were identified through data sharing with collaborators and screening databases of several diagnostic and research genetic laboratories worldwide, as well as using GeneMatcher^16^. Informed consent forms allowing for participation were signed by all study participants and/or their parents or guardians. Genome/exome sequencing (GS/ES) was performed on genomic DNA extracted from blood in different diagnostic or research laboratories worldwide, and if required, candidate variants were confirmed by Sanger sequencing in the available samples from other members of the families.

#### Clinical assessment

Detailed clinical data and family history were collected for new and reported cases in the form of completing a clinical proforma by the recruiting clinicians. Brain MRIs were reviewed by an experienced pediatric neuroradiologist (M.S.). Video segments of seven patients were suitable for fine analysis of the stereotypies by an experienced neurologist (E.F.). Facial photographs and/or videos were reviewed for 24 patients from 15 families, including 18 new patients from 11 families and 6 previously published patients from 4 families^12–14^. Their dysmorphic features were described based on the terminology recommended by Elements of Morphology^17^. Where no term was available for a dysmorphic feature seen in a patient, HPO terminology was used instead^18^.

### Drosophila studies

#### Drosophila husbandry

All stocks and experimental crosses were raised on standard fly-food media and kept at 25 °C with 12 h light: 12 h dark cycles. *Drosophila* strains used in this study are listed in Supplementary Table 1. For behavioural experiments, isogenised lines (indicated in Supplementary Table 1) were generated by outcrossing each mutation or transgene insertion into the iso31 strain of *w^11^*^18^ for five generations^19^.

#### Immuno-histochemistry

Immunohistochemical experiments were performed as previously described^20^. Briefly, adult or larval brains were dissected in PBS, and fixed in 4% paraformaldehyde (MP biomedicals) for 20 min at room temperature. Tissues were washed with PBST (PBS, 0.3% Triton X-100), blocked in 1% goat serum in PBST and incubated in primary antibody overnight at 4 °C. Following primary antibody incubation, tissues were washed a further three times in PBST and incubated overnight in secondary antibody. Antibodies used in this study include mouse anti-Elav (Developmental studies hybridoma bank – Elav-9F8A9)^21^, rabbit anti-cleaved DCP1 (Cell signalling technology – #9578), and mouse anti-Repo (Developmental studies hybridoma bank – 8D12)^22^.

#### *Drosophila* behavioural analyses

*Drosophila* activity was assayed using the *Drosophila* Activity Monitor system (DAM; Trikinetics, MA, USA) as previously described^23,24^. Briefly, individual flies obtained between 3-5 days after eclosing were loaded into glass tubes containing 4% sucrose and 2% agar (w/v) and sealed with cotton-wool plugs. Monitors were kept at 25°C with 12 h light: dark cycles for two days to acclimatise. On the third day activity was recorded for 24 h. For measurements of peak activity at ZT0-1 or ZT12-13 (ZT: zeitgeber), activity was taken from the hour after lights on or off during the third day. DAM data was analysed using the Rethomics R package^25^. Only flies surviving for the full three days were included for analysis. For adult-specific knockdown and rescue experiments, flies were raised at 18°C until 2 days post-eclosion, at which point they were loaded into DAM monitors and moved to 29°C for 3 days (or remained at 18°C for controls).

Larval locomotion assays were conducted by transferring wandering 3^rd^ instar larvae to a large arena containing 2% agar. The arena was placed into a 25°C incubator and larvae were left to acclimatise for 30 s. Larval crawling was video recorded for 1 min. Video files were analysed using ImageJ to calculate total distance travelled.

Negative geotaxis (climbing) assays were conducted as previously described^26^. Briefly, cohorts of 10 flies were transferred to clean glass measuring cylinders and left to acclimatise for at least 20 mins. Flies were firmly tapped down 3-5 times and number of flies crossing an 8 cm vertical threshold in 12 s was recorded. Three technical replicates were included for each genotype.

#### Statistical analyses

Statistical data analysis was performed using R or GraphPad Prism. Datasets were first tested for normality using the Shapiro-Wilk normality test. Statistical analyses were performed using a t-test with Welch’s correction or one-way ANOVA with Dunnett’s multiple comparisons post-hoc test if data were normally distributed; and Mann-Whitney U-test or Kruskall-Wallace test with Dunn’s multiple correction if data were non-normally distributed.

## Results

### Clinical profile of the study cohort

The overall cohort comprises 13 females and 15 males, whose age at last evaluation ranged between 2 and 36 years (median 11, IQR 11). An overview of the clinical findings can be found in Figure 1A and Table 1. Detailed clinical information is available in Supplementary Table 2. Consanguinity was reported in 15 families (83%). Pregnancy and delivery were unremarkable for all the patients for whom information was available (24/24, 100%) and all newborns were at term. Birth parameters of length and weight, when available, were within normal ranges for all the infants, and only one presented with decreased head circumference at birth (HP:0011451). Most of the newborns (26/28, 93%) manifested infantile hypotonia (HP:0008947). Failure to thrive (HP:0001508) and feeding difficulties in the infantile period were documented in 36% and 37% of those examined, respectively (5/14 and 7/19).

**Figure 1.**
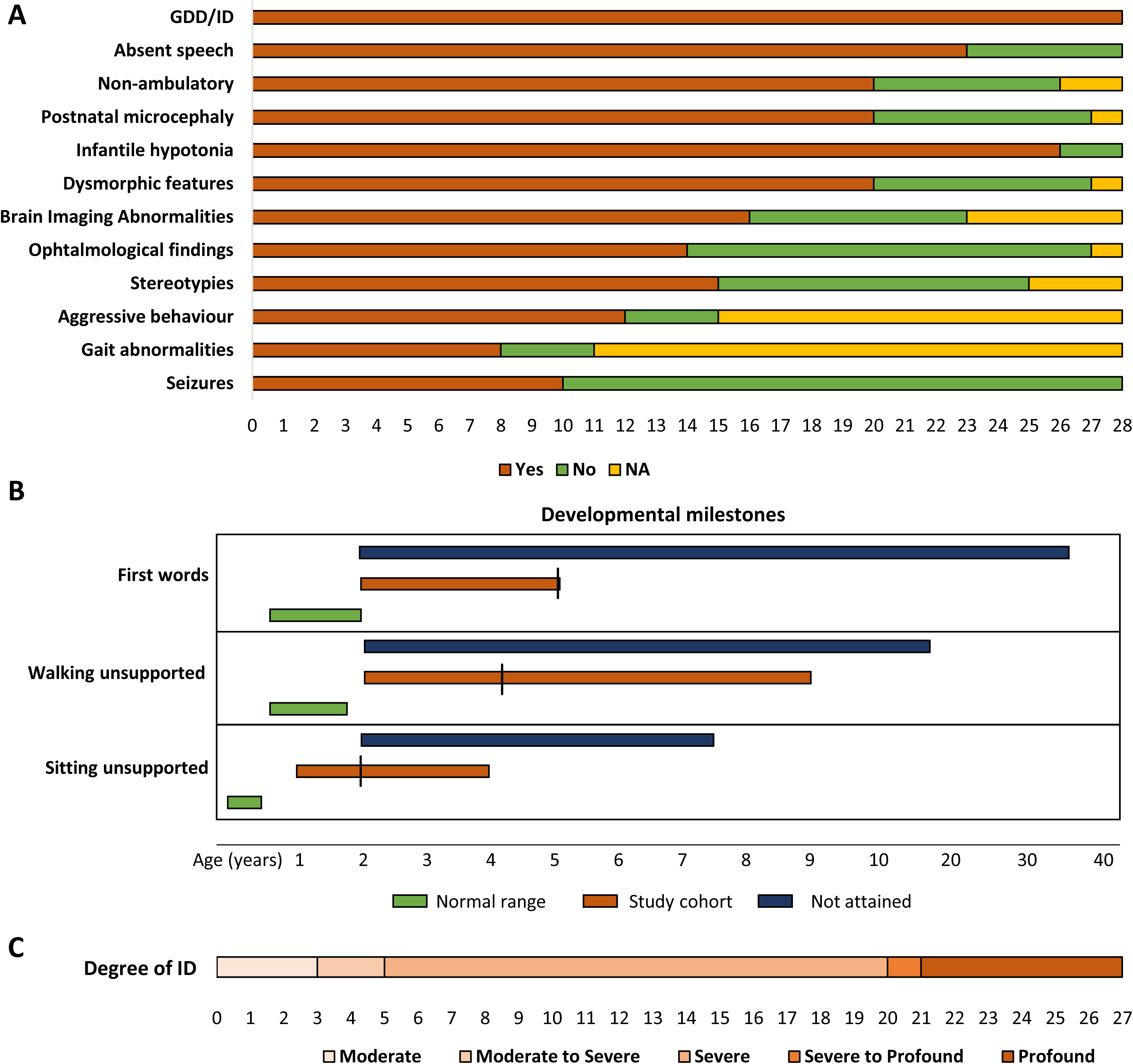
*RBL2-*related disorder is characterized by a range of neurological, behavioural and developmental abnormalities. **A**. Representation of the most frequent clinical features observed in the *RBL2* patients (Y axis: clinical features, X axis: number of patients). **B.** Timeline-style schematic outlining the acquisition of key developmental milestones observed in the affected individuals. Most of the individuals did not attain independent sitting, walking or speech development (blue bar indicates range at last evaluation), while the others presented delayed acquisition (orange, line indicates median age). Normal range is indicated in green. **C.** Schematic depiction of the degree of intellectual disability observed in the patients (number of patients indicated in bottom line). The spectrum ranged from moderate (left) to profound (right).

**Table 1.**
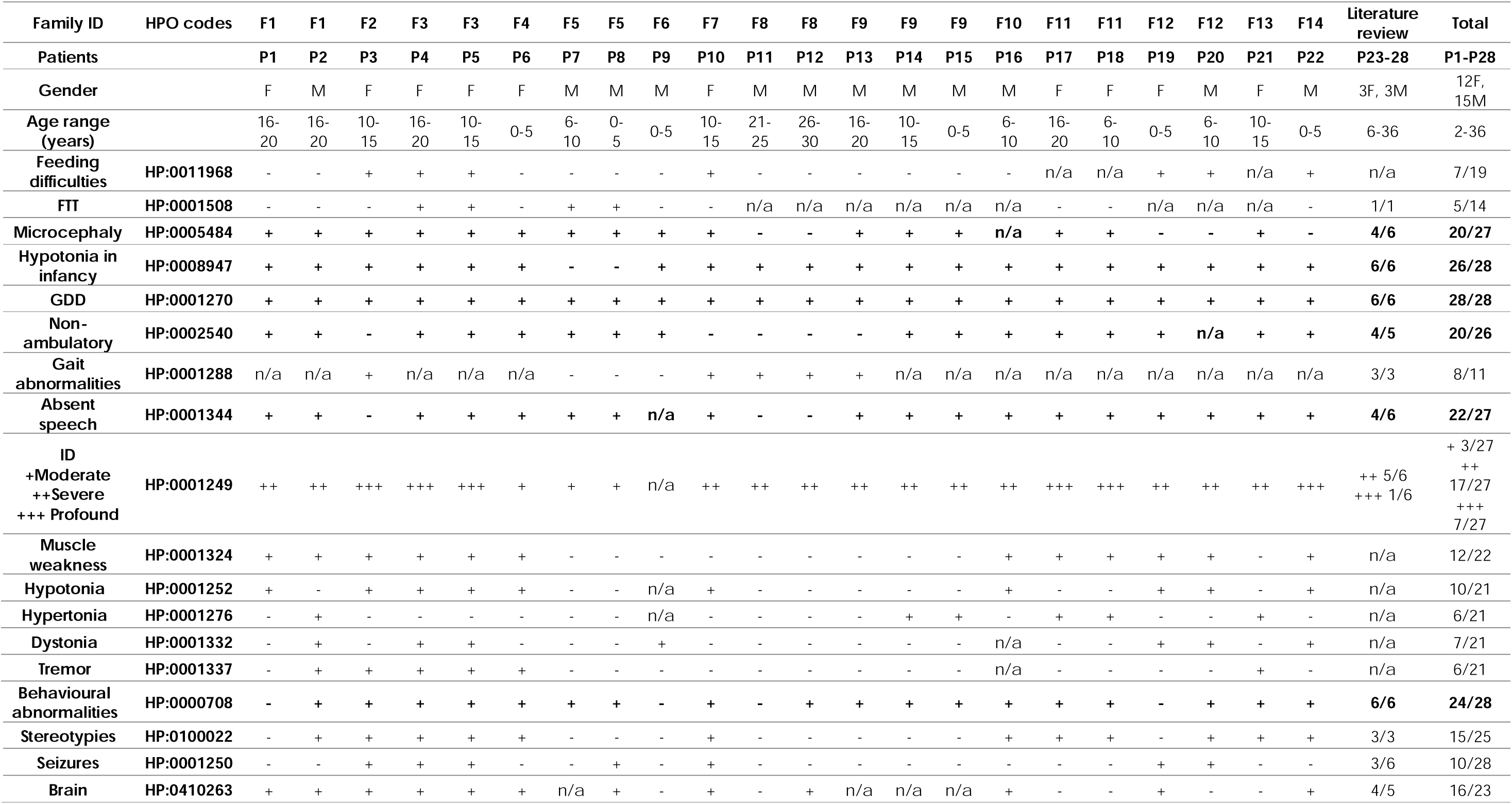
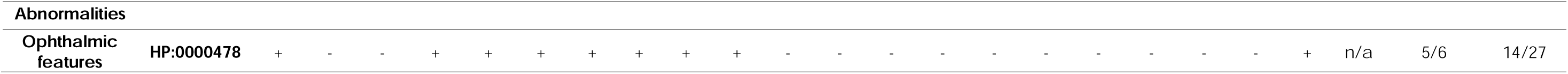
Overview of the clinical features observed in the cohort.

Global developmental delay (HP:0001263) and intellectual disability (HP:0001249) were reported in all the affected individuals (28/28, 100%). All patients (28/28, 100%) presented motor delay (HP:0001270): while most of the affected children only presented a delay in the achievement of unsupported sitting (21/23 delayed, 2/23 not attained). The majority never attained independent walking (19/27, 70%) and the remainder had delayed acquisition. In the same way, most children (22/27, 81%) showed complete lack of development of speech and expressive language abilities (HP:0001344), while in the remaining individuals (5/27, 19%) development of speech was delayed (HP:0000750) and involved the use of just a few words (Figure 1B). Regression of motor and cognitive abilities (HP:0002376) was reported in three patients (3/22, 14%). When formal cognitive assessment was performed, degree of intellectual disability ranged from moderate (3/27), severe (17/27), to profound (7/27) (Figure 1C). Behavioural abnormalities (24/28, 86%) included stereotypies (15/25, 60%), aggressive behaviour (12/15, 80%), autism (6/21, 28%) and, when data were available, sleep disturbance (5/5). Video segments of seven patients were suitable for fine analysis of the stereotypies. The stereotypies usually involved the cervico-facial area (head and/or orofacial region) along with the distal part of the upper limbs, typically in the form of hand clasping/squeezing and mouthing, and finger wringing (Video 1). Seizures occurred in 36% of the individuals (10/28). Age of onset of the seizures ranged from 1 to 20 years (median age 8 years, IQR 7). Patients mostly presented a generalized seizure(HP:0002197) (n=6) onset, while focal onset (HP:0007359) occurred in two patients, one with secondary generalization. Seizure types were variable and included tonic-clonic (n=3), myoclonic (n=2), and tonic (n=1). Where EEG was available, abnormalities were documented in 7/10 patients examined (70%), including one patient with no clinical seizures, and included focal, multifocal, and diffuse epileptiform discharges, slowing of background activity and subcortical changes.

Neurological examination showed increased tendon reflexes (HP:0001347) (10/17, 59%), muscle weakness (HP:0001324) (12/22, 55%), axial hypotonia (HP:0008936) (10/21, 47%) and spasticity (HP:0001257) (11/25,44%). Ophthalmological evaluation revealed presence of abnormal findings in half of the cases (14/27, 52%), including strabismus (HP:0000486) (7/19, 37%), nystagmus (HP:0000639) (6/26, 23%), refractive defects (3/25,12%), poor vision (6/26, 23%), optic disc anomalies (4/24, 17%) and orbital mass (2/26, 8%).

At last evaluation, 74% of the patients (20/27) were microcephalic (HP:0000252) (Figure 2A). Dysmorphic features were described in 70% of the cases and included, based on photographic assessment, low anterior hairline (50%), narrow forehead/bifrontal/bitemporal narrowing (83.3%), full or broad nasal tip (77.8%), thick/full lower lip vermilion (66.7%) and broad or tall pointed chin (77.8%) (Figure 2B, Supplementary Table 3).

**Figure 2.**
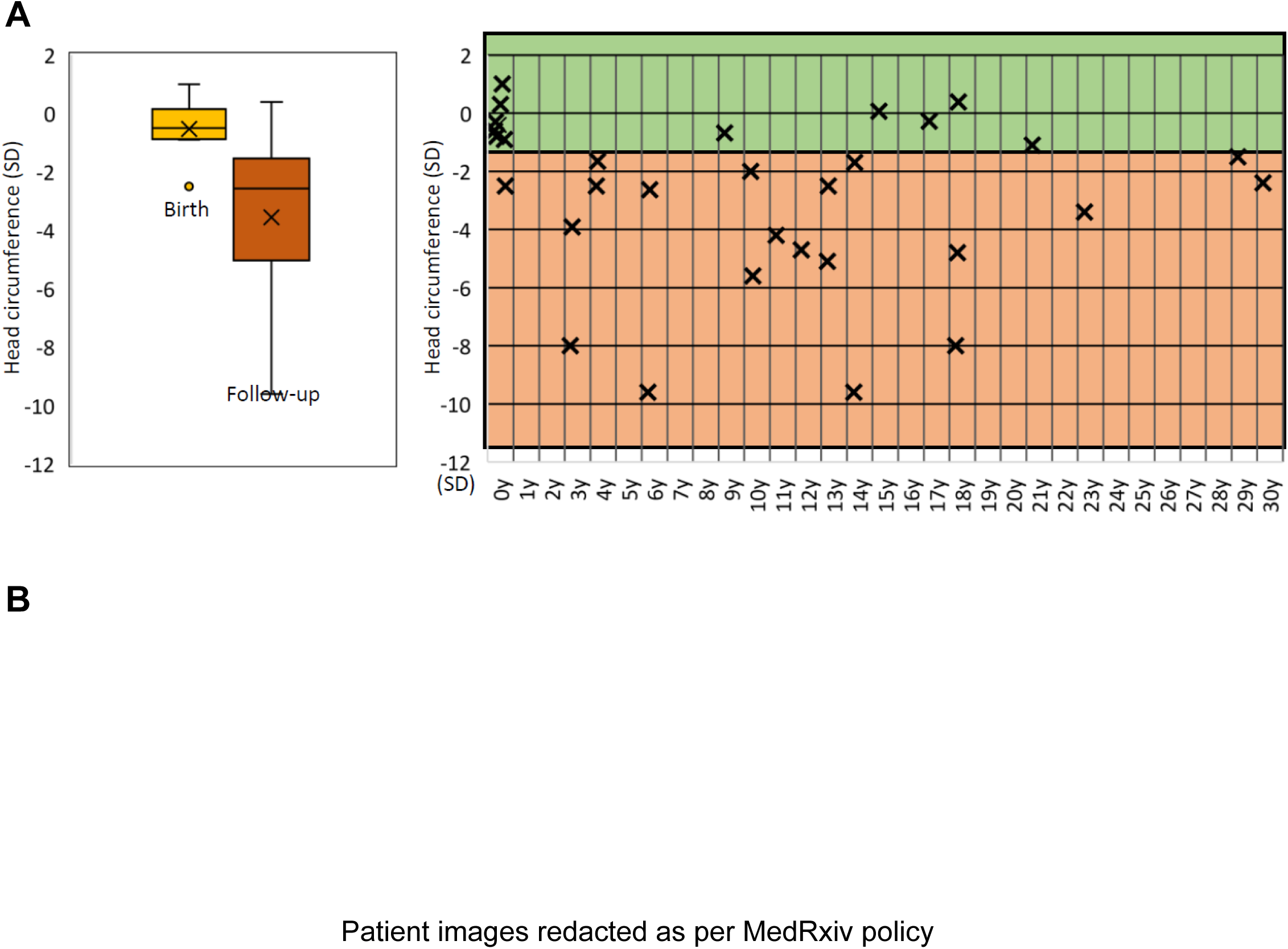
*RBL2* patients present postnatal microcephaly and dysmorphic features, without a recognizable facial ‘gestalt’. **A**. Schematic representation of head circumference measurements, expressed in standard deviations (SD). Head circumference was within normal ranges at birth, while reduced at last examination (left panel). The right panel illustrates age at last follow-up and measurements in SD. **B.** Facial features of the patients.

### Neuroradiological features of *RBL2* patients

Brain MRIs were available for review in 12/28 cases (mean age at MRI 7 years, range 8 months – 17 years). The most frequent neuroimaging finding was a mild-to-moderate cerebral atrophy with antero-posterior gradient and thin corpus callosum (10/12, 83.3%) (Figure 3). Reduced white matter volume with ventricular enlargement was associated in 8/10 cases. In 9 subjects (75%), we found white matter signal abnormalities, including faint to marked focal signal changes at the level of forceps minor (8/9), delayed myelination (2/9), and multiple patchy frontal signal changes (1/9). Mild-to-moderate cerebellar atrophy was noted in 7/12 individuals (58.3%), with dentate signal changes in 3 cases and clear progression in one subject with a follow-up MRI; in one individual there were also bilateral widespread subcortical signal changes. In 4 other subjects (33%) there was hypoplasia of the inferior portion of the cerebellar hemispheres and/or vermis, with associated foliar anomalies in 1 case. Optic nerve thinning was detected in 5/12 (41%) individuals. Calcifications in the basal ganglia were found in 2/12 (16.6%) cases. Finally, expansile lesions were found in 2 subjects: a large mass extending from the III ventricular floor to the prepontine cisterns (hypothalamic hamartoma versus ectopic cerebellar tissue) in P6 and a cystic mandibular lesion in P23 (Supplementary Table 4).

**Figure 3.**
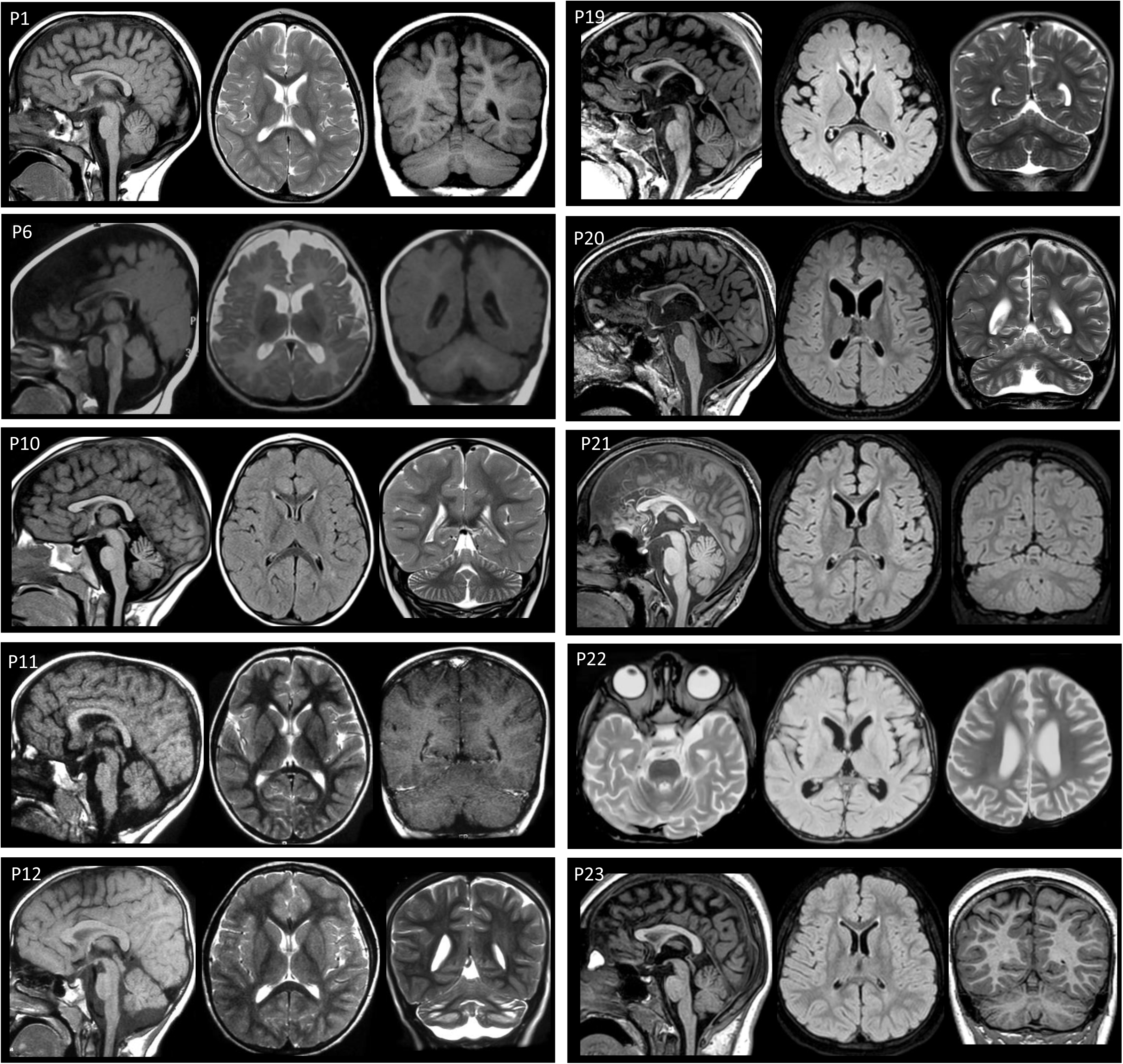
Molecular spectrum of loss-of-function variants in *RBL2.* **A**. Schematic representation of variants location on the *RBL2* gene. Upper part: newly reported variants. Lower part: previously reported variants. **B.** Pedigrees of the newly reported patients. Solid black, affected. Genotype, where indicated, represent results of segregation. **C.** Classification of variants according to type.

### Molecular spectrum of *RBL2* variants

A total of 19 *RBL2* variants are included in this study (Figure 4A), 14 of which are newly reported variants not described in the literature. Within the cohort of newly reported families (Figure 3B), only one affected family carried a previously reported variant (c.556C>T, p.Arg186Ter). Molecular findings are shown schematically in Figure 4 and described in detail in Supplementary Table 5. The variants were inherited from unaffected heterozygous parents: 23 patients inherited the variant in the homozygous state and 4 in compound heterozygous state. All variants were either absent or found at very low allele frequencies in multiple variant frequency databases (range 0.0-0.00002). The molecular spectrum hereby described includes truncating (n=5), frameshift (n=6) splice (n=6) and large deletions (n=2) (Figure 4C).

**Figure 4.**
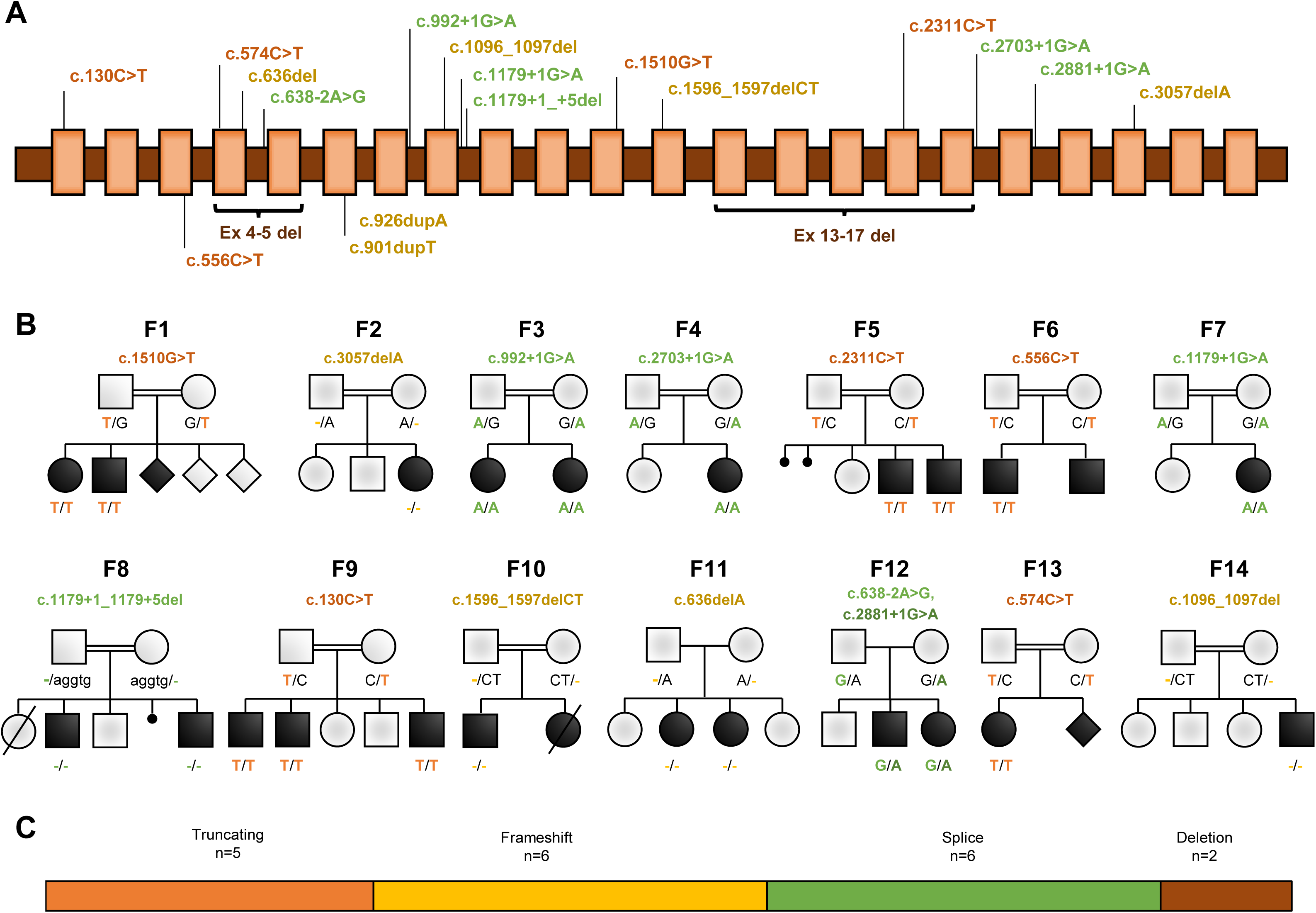
Neuroimaging features of RBL2-related disorder. Sagittal T1-weighted image (first image), axial T2-weighted or FLAIR image (central image), coronal T1 or T2-weighted or FLAIR image (last image). Most subjects have an enlargement of the cerebral CSF spaces with an antero-posterior gradient associated with thinning of the corpus callosum, particularly in the anterior portions. There is additional cerebellar atrophy in P10, P12, P20, P22, and P23. Bilateral mild-to-moderate signal changes are noted at the level of the forceps minor in P6, P19, P20, P21, P22, and P23. Note the large prepontine lesion in P10.

According to the American College of Medical Genetics (ACMG) classification, six were classified as pathogenic and thirteen as likely pathogenic. All identified variants were predicted as damaging across a suite of in silico tools and expected to lead to LOF of the protein. Our findings therefore confirm a clear link between biallelic pLOF mutations in *RBL2* and a multifaceted neurodevelopmental disorder.

### A *Drosophila* model of RBL2-linked pathology recapitulates morphological patient phenotypes

The *Drosophila melanogaster* genome encodes two Rb proteins: Rbf and Rbf2. Of these, Rbf shares the greater similarity to RBL2 (39% similarity and 25% identity; compared to 35% similarity and 20% identity for Rbf2). Indeed, fourteen distinct databases of orthology relationships place Rbf as the closest *Drosophila* orthologue of RBL2, and RBL2 was the closest match for Rbf in a reverse orthology search (https://flybase.org/reports/FBgn0015799#orthologs).

Similar to RBL2, prior work has shown *Drosophila* Rbf interacts with and negatively regulates E2F transcription factor activity to repress cell-cycle gene expression^27–29^. Thus, human RBL2 and *Drosophila* Rbf exhibit functional as well as amino-acid conservation. Published single-cell RNAseq data further indicate that *Rbf* is widely expressed throughout the *Drosophila* nervous system, whereas *Rbf2* is not (Supplementary Fig. 1A, B)^30–31^. Hence, we investigated how loss of Rbf function impacted neural development and behaviour in *Drosophila*.

We first set out to determine the extent to which *Drosophila Rbf* loss of function phenotypes resemble *RBL2* patient symptoms. Initially, we examined male and female flies hemizygous or homozygous respectively for a hypomorphic allele of *Rbf* (*Rbf^120a^*) to determine whether partial loss of Rbf function in flies recapitulated morphological and behavioural phenotypes observed in *RBL2* patients. While a previous study suggested that eye morphology in *Rbf^120a^* hemizygotes was relatively normal^32^, we noticed that the size of the eye was significantly smaller in hemizygous *Rbf^120a^* males compared to control flies, although the highly organised ommatidial structure appeared unaffected (Figure 5A, B). We also examined the eye-size of female *Rbf^120a^* homozygotes and females trans-heterozygote for *Rbf^120a^* and the *Rbf*^14^ null allele (note that adult *Rbf*^14^ homozygotes are embryonic lethal)^33^. Both *Rbf^120a^* homozygote and *Rbf^120a^/Rbf^14^* trans-heterozygous females also displayed smaller eyes compared to wild-type control and *Rbf^120a^/+* or *Rbf^14^/+* heterozygote flies (Figure 5C).

**Figure 5.**
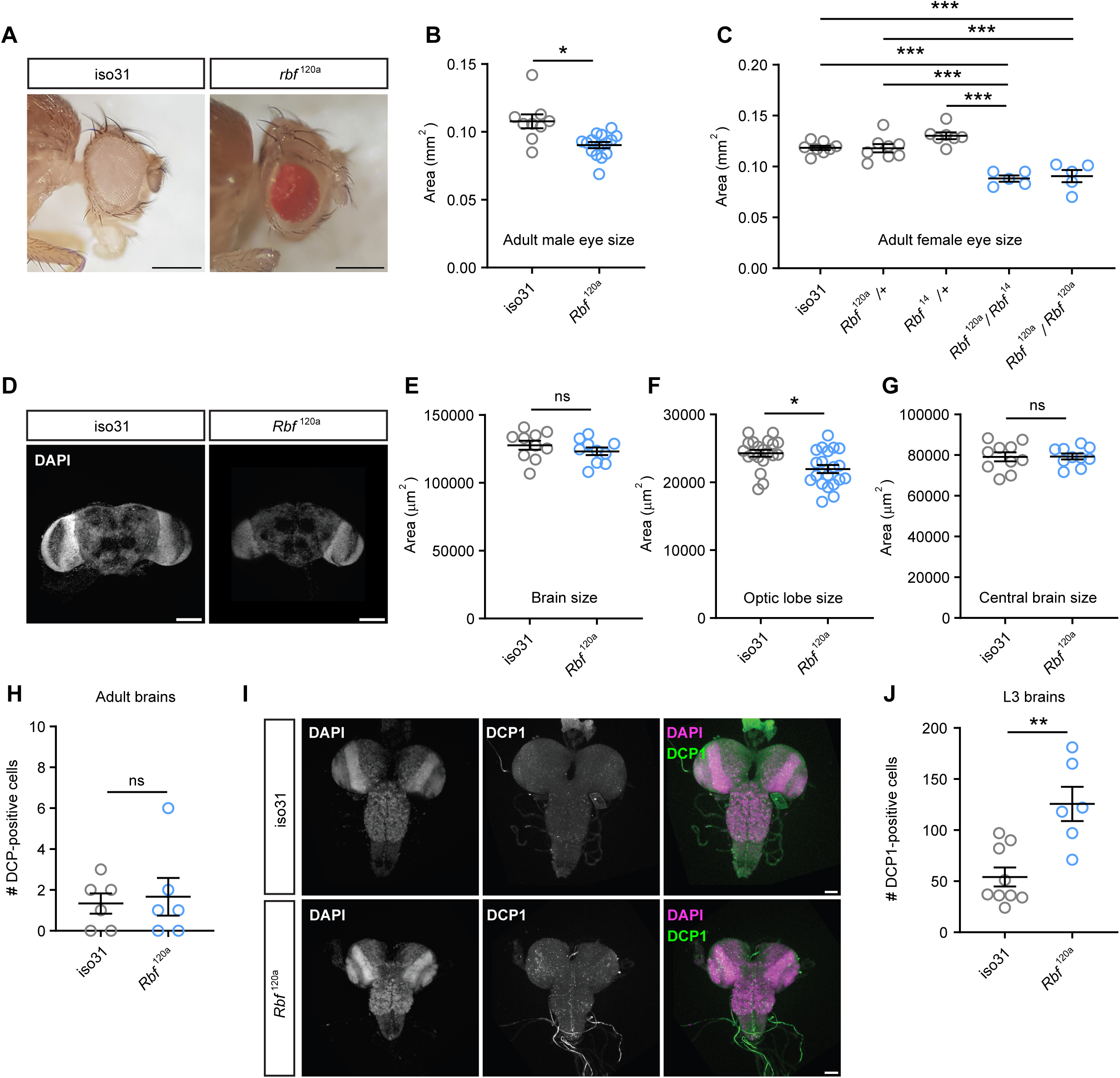
Loss of Rbf activity reduces neuronal growth in *Drosophila*. **A**. Representative images of adult eyes in control (iso31) and *Rbf^120a^* hypomorphs. Scale bars = 0.3 mm. **B**. Quantification of eye sizes in male *Rbf^120a^* hemizygotes (n = 16) compared to controls (n = 9). **C.** Quantification of eye sizes in female *Rbf* allelic combinations (n = 5-8). **D**. Representative images of adult brains in control and *Rbf^120a^* adult males. Scale bars = 50 μm. **E-G.** Measurements of brain morphology in control and *Rbf^120a^* hemizygotes adult males (n = 10 brains, 10 central brains, and 20 optic lobes, per genotype). **H**. Quantification of apoptotic (DCP1 positive) cells in control and *Rbf^120a^* hemizygotes adult male brains (n = 6 per genotype). **I**. Representative images of DCP1-lablled control (n = 9) and *Rbf^120a^* hemizygote (n = 6) third instar larval nervous system. Nuclei are counterstained with DAPI. Scale bars = 20 μm. **J.** Quantification of apoptosis in control and *Rbf^120a^*hemizygotes third instar larval brains. Error bars: SEM. * p< 0.05, ** p<0.005, *** p< 0.0005, ns – p> 0.05, unpaired t-test with Welch’s correction (B, E, F), one-way ANOVA with Dunnett post-doc test (C), Mann-Whitney U-test (G, H, J).

Since microcephaly is a clinical feature of *RBL2* patients, we next examined whether brain size was also reduced in *Drosophila Rbf* mutants (Figure 5D-G). While overall brain size was not significantly smaller in *Rbf^120a^* hemizygote males (Figure 5E), we observed a significant reduction in the size of *Rbf^120a^* hemizygote optic lobes, visual processing centres that contain > 60% of all neurons in the fly brain^34^ (Figure 5F). In contrast, the central brain region of *Rbf^120a^* hemizygotes was unaltered (Figure 5G).

Rb proteins have been linked to apoptosis in human^35^ and *Drosophila*^36^, with *Rbf^120a^* mutants displaying increased apoptosis in the eye imaginal disc (the developmental precursor to the adult eye^32^). We therefore reasoned that decreased brain size in *Rbf* mutants might be driven by an increase in cell death. To determine the amount of apoptosis in the brains of *Rbf^120a^* mutants, we stained tissues with anti-DCP1, which recognises the cleaved version of a caspase protein involved in apoptotic cell death. Examination of adult *Rbf^120a^*brains indicated minimal apoptosis, as was similarly observed in control adult brains (Figure 5H). However, examination of larval brains, in which most neurons are in a more immature state, revealed significantly greater numbers of apoptotic cells in *Rbf^120a^*mutants than controls (Figure 5I, J). This suggests that neuronal precursors and immature neurons are more sensitive to the induction of apoptosis when Rbf is depleted, in agreement with previous observations of the developing eye^32^. Overall, the morphological phenotypes we observe in the adult head and brain are consistent with the microcephaly seen in *RBL2* patients, indicating a conserved function for the RBL2 and Rbf proteins in controlling head morphology during development.

### *Drosophila Rbf* mutants display locomotor defects

Since profound motor delay was observed in all patients homozygous for *RBL2* variants, we also tested whether *Drosophila Rbf* mutants exhibited motor defects. To do so, we utilised the *Drosophila* Activity Monitor (DAM) system^23^, which quantifies spontaneous activity by recording the number of times individual flies interrupt an infra-red beam bisecting a glass tube housing each fly (Figure 6A). *Rbf^120a^* hemizygotes showed significantly lower locomotor activity compared to controls both over a 12 h light: 12 h dark period (Figure 6B) and during a one-hour window following lights-on (ZT0-1; ZT – zeitgeber time) that corresponds to a period of peak activity (Figure 6C). We observed a similar effect in female *Rbf^120a^* homozygotes and *Rbf^120a^/Rbf^14^*trans-heterozygotes, but not in females that are heterozygous for either allele (Figure 6D), thus confirming that the above alterations in locomotor activity were caused by mutations in *Rbf*. To further characterise these behavioural abnormalities, we conducted negative geotaxis (climbing) assays^26^. *Rbf^120a^* hemizygote males displayed significantly lower climbing ability compared to control animals (Supplementary Figure 1C, D), further indicating that *Rbf* LOF induces significant motor defects in flies. Thus, *Drosophila Rbf* mutants recapitulate morphological and behavioural phenotypes observed in patients harbouring *RBL2* mutations.

**Figure 6.**
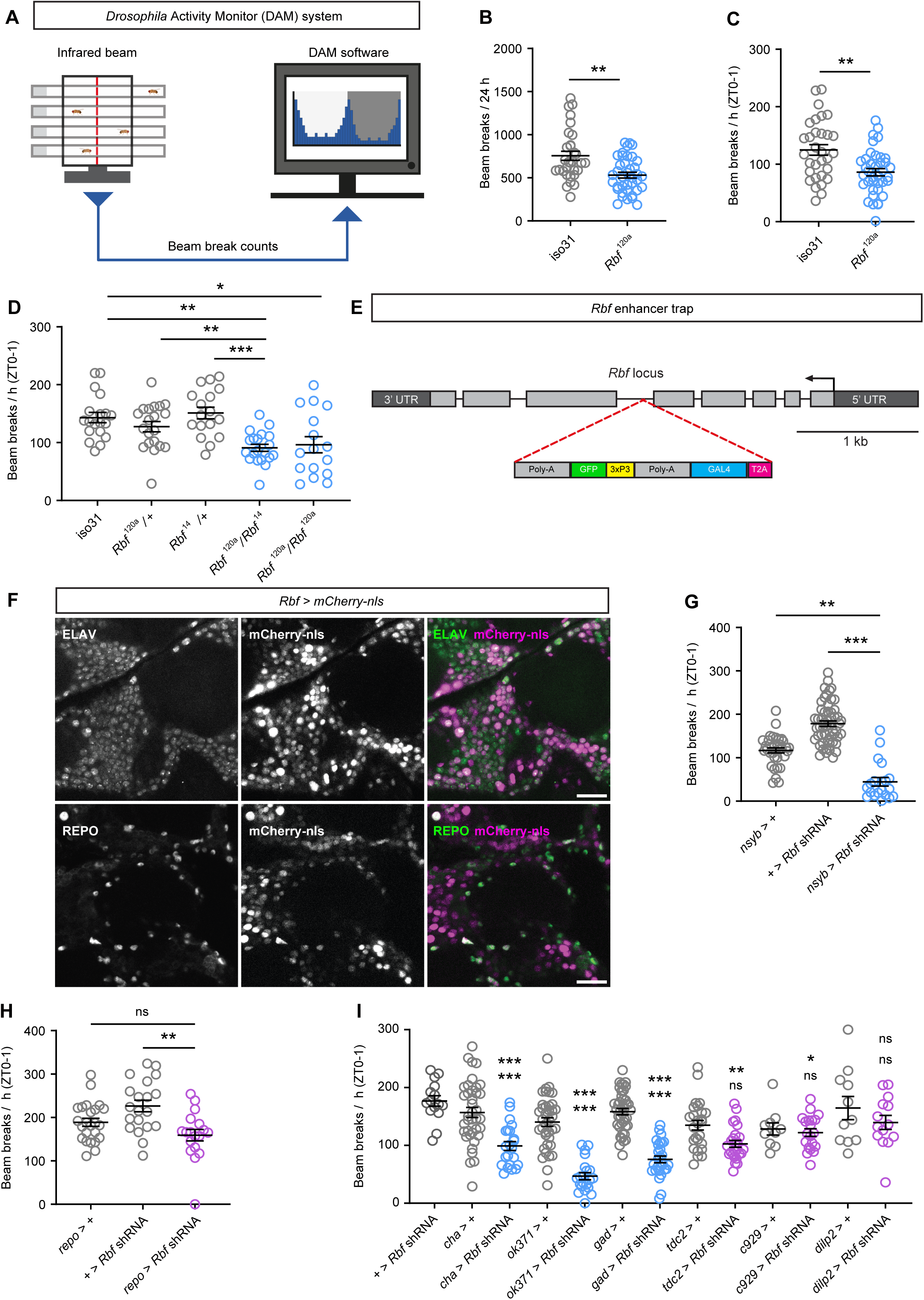
Rbf acts in post-mitotic neurons to promote movement in *Drosophila*. **A**. Schematic representation of the *Drosophila* Activity Monitor (DAM) system. **B-C**. DAM activity in *Rbf^120a^* hemizygotes (n = 38) and controls (iso31; n = 31) across a 24 h period (B) or during zeitgeber time (ZT) 0-1, a period of peak activity (C). **D.** DAM activity in adult females harbouring trans-heterozygote or heterozygote *Rbf* allelic combinations, and wild type iso31 controls during ZT0-1. n = 16-20. **E.** Schematic representation of the CRIMIC insertion in the *Rbf* locus, which allows for *Rbf*-dependent Gal4 reporter expression. **F.** *Rbf*-Gal4 driven nuclear mCherry expression in the adult central brain. Neurons and glia are counterstained with antibodies against ELAV and REPO respectively. Scale bar = 20 µm. **G.** Pan-neuronal post-mitotic knockdown of *Rbf* severely reduces peak locomotor activity during ZT0-1. n = 20-54. **H.** Knockdown of *Rbf* in glial cells (using *repo*-Gal4 to express *Rbf* shRNA) does not significantly reduce peak locomotor activity during ZT0-1 compared to both driver and transgene alone controls (n = 18-24). **I.** Knockdown of *Rbf* in cholinergic, glutamatergic, and GABAergic neurons, reduces peak activity during ZT0-1 in adult males. n = 11-41. Upper significance notation is relative to *Rbf* shRNA alone controls, lower significance notation is relative to Gal4 driver alone controls. Error bars: SEM. * p< 0.05, ** p<0.005, *** p< 0.0005, ns – p> 0.05, unpaired t-test with Welch’s correction (B), Mann-Whitney U-test (C), one-way ANOVA with Dunnett post-doc test (D, H, I), or Kruskal-Wallis test with Dunn’s post-hoc test (G).

### *Rbf* is highly expressed in adult neurons

Given the strong phenotypic similarities between humans and fruit flies harbouring *RBL2/Rbf* LOF mutations, we tested whether we could utilise *Drosophila* to probe the mechanistic basis of *RBL2*-linked neurodevelopmental defects. Rb proteins are well-known for their role in transcriptional repression of cell-cycle related genes at the G1/S phase transition^37^. Hence, it is expected that *Rbf* would be expressed in the developing brain. However, it is unclear whether Rbf also continues to play a role in fully differentiated neurons following cell-cycle exit. To investigate which cells in the nervous system express *Rbf*, we took advantage of a CRISPR-mediated insertion of a Gal4 cassette (CRIMIC insertion) in an *Rbf* intron, which results in expression of Gal4 under control of *Rbf* regulatory sequences (termed *Rbf-*Gal4 hereafter)^38^ (Figure 6E). Crossing these flies to a UAS*-mCherry-*nls line yields expression of nuclear mCherry as a reporter of *Rbf* expression. Examination of *Rbf-*Gal4 activity in larval brains revealed widespread expression, indicating that *Rbf* is indeed broadly expressed in the developing brain (Supplementary Figure 1E). More surprisingly, adult brains – which do not display appreciable neurogenesis under normal conditions – also exhibited widespread *Rbf*-driven mCherry expression that co-localised with the neuronal marker Elav (Figure 6F and Supplementary Figure 1F). Hence, *Rbf* expression persists in neurons long after terminal cell-cycle exit. In contrast, only a small population of Repo-labelled glial cells expressed mCherry under the control of *Rbf*-Gal4 (Figure 6F). These findings are consistent with published single-cell RNAseq data showing that *Rbf* is preferentially expressed in post-mitotic neurons relative to glia (Supplemental Figure 1A).

### Rbf knockdown in neurons causes severe behavioural defects

To directly test for an underappreciated role of Rbf in fully differentiated post-mitotic cells, we examined whether reducing Rbf expression in post-mitotic neurons resulted in locomotor defects similar to those observed in constitutive *Rbf* hypomorph flies. To do so, we used transgenic RNAi to deplete *Rbf* specifically in fully differentiated neurons using the *nSyb*-Gal4 driver. Strikingly, using the DAM system once more, we found that pan-neuronal knockdown of *Rbf* with a previously verified shRNA-expressing line^39^ severely reduced peak movement in adult flies (Figure 6G). To rule out off-target effects, we repeated these experiments using two additional RNAi lines targeting *Rbf* mRNA. Both constructs similarly reduced peak movement when expressed in post-mitotic neurons (Supplementary Figure 2A, B). In contrast to neuronal knockdowns, RNAi-mediated depletion of *Rbf* in glial cells did not significantly reduce peak locomotor activity (Figure 6H), in accordance with the above observation that *Rbf* expression is less abundant in glia than neurons (Figure 6F).

We next used climbing assays to quantify stimulus-induced negative geotaxis. These assays further indicated that flies with reduced Rbf expression in neurons have severe motor defects, showing significantly reduced climbing ability compared to controls (Supplementary Figure 2C). Interestingly, larval locomotion was unchanged in either *Rbf* hypomorphs or following knockdown of *Rbf* in post-mitotic neurons, suggesting that larval neuronal lineages have a differential requirement for Rbf compared to their adult counterparts (Supplementary Figure 2D, E). Importantly, *Rbf* knockdown in adult post-mitotic neurons did not reduce optic lobe size nor induce a measurable increase in neuronal apoptosis (Supplementary Figure 2F, G). Taken together, these data suggest that *Rbf* plays important neuron-autonomous roles that are essential for adult locomotor behaviour and which are independent of neuronal viability.

### Multiple neuronal subtypes are affected by Rbf knockdown

To identify which cell-types in the post-mitotic brain are affected by *Rbf* knockdown, we used specific drivers to restrict *Rbf* shRNA expression to genetically defined subsets of neurons. We knocked down *Rbf* in discrete neuronal subtypes, including cholinergic, GABAergic, and glutamatergic neurons. Of these, *Rbf* knockdown in glutamatergic neurons (which include *Drosophila* motoneurons) yielded the most significant decline in locomotor activity, as measured using the DAM system (Figure 6I and Supplementary Figure 3). *Rbf* knockdown in GABAergic and cholinergic neurons did not significantly decrease overall activity across 24 h (Supplementary Figure 3). However, in the one-hour period following lights-on (ZT0-1), during which control flies exhibit a peak period of locomotor activity, both cholinergic and GABAergic *Rbf* knockdown flies showed significantly reduced activity (Figure 6I), indicating a partial perturbation of locomotor capacity. We further tested the motor defect of these flies by conducting climbing assays. These experiments confirmed that reduced *Rbf* expression in glutamatergic, cholinergic, or GABAergic neurons, resulted in significantly decreased climbing ability compared to controls (Supplementary Figure 2C). In contrast, *Rbf* knockdown in peptidergic neurons did not perturb overall or peak locomotor activity (Figure 6I and Supplementary Figure 3). These data demonstrate that Rbf acts in several post-mitotic neuronal subtypes to modulate movement, with glutamatergic neurons representing a particularly relevant cellular locus.

### Post-mitotic restoration of Rbf rescues locomotor defects in *Rbf* hypomorphs

Since *Rbf* mutants display morphological phenotypes consistent with cell-cycle defects and apoptosis during development, but also behavioural abnormalities that can be induced by knockdown of *Rbf* in post-mitotic neurons, we questioned whether locomotor phenotypes in constitutive *Rbf* hypomorphs were due to developmental defects or reduced *Rbf* expression post-neurogenesis (i.e. in post-mitotic neurons). To address this question, we expressed Rbf solely in fully differentiated neurons in the *Rbf* hypomorph background. Interestingly, this manipulation fully rescued the reduced peak activity of *Rbf^120a^*hypomorphs, while over-expression of *Rbf* in a wild-type background had no effect on peak locomotor activity (Figure 7A).

**Figure 7.**
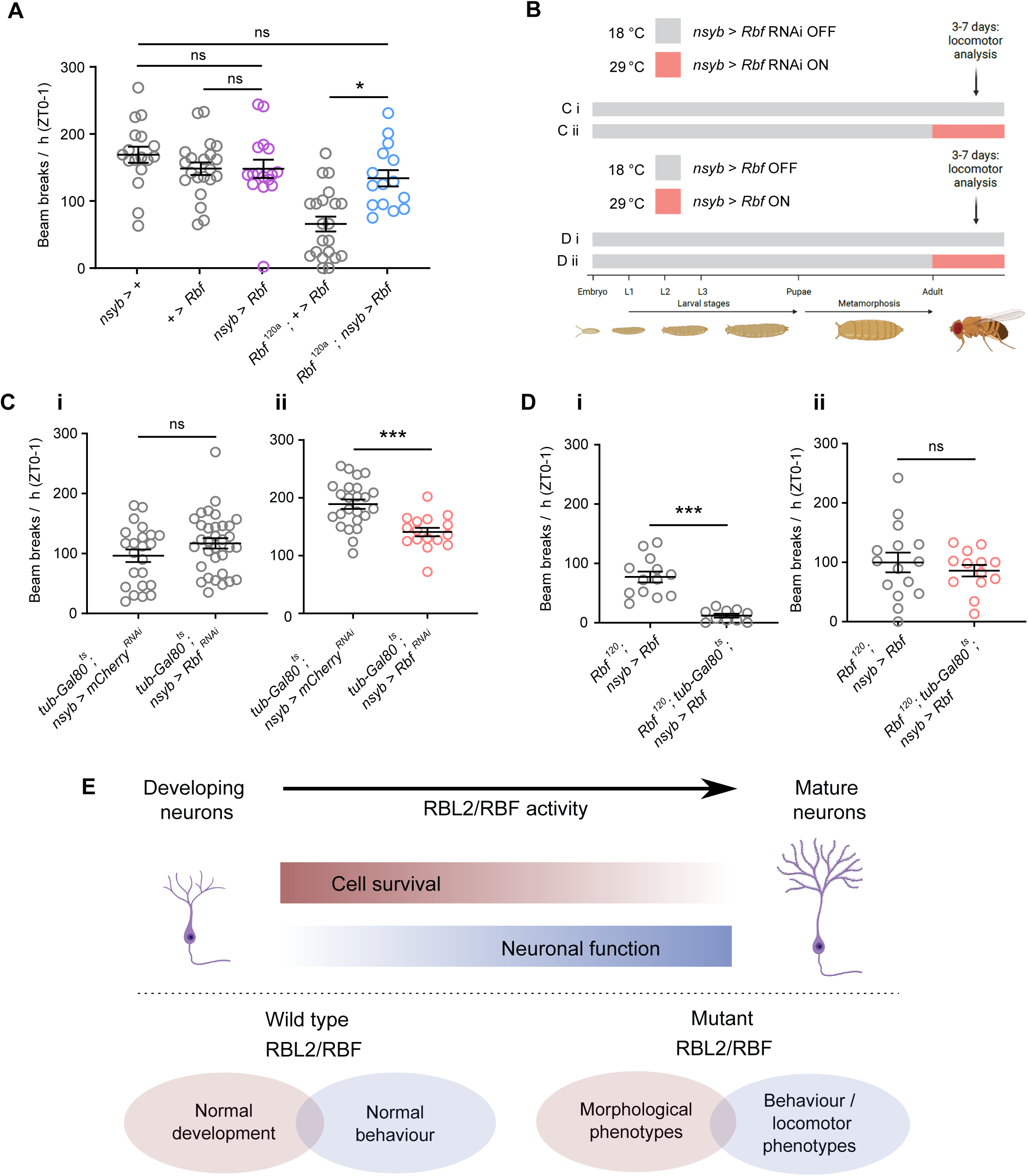
Adult-stage neuronal expression of Rbf rescues locomotor defects in *Rbf* hypomorphs. **A**. Effects of post-mitotic, neuron-specific *Rbf* expression on peak locomotor activity in either wild-type or *Rbf^120a^* hypomorph backgrounds. Data are from adult males. n = 15-21. **B.** Experimental paradigm for temperature-induced knockdown and rescue experiments shown in panels C and D. **C.** Quantification of peak activity for control adult male flies kept at (i) non-permissive temperature: *mCherry* (n = 33) or *Rbf* (n = 23) shRNA expression – repressed; and (ii) experimental adult male flies maintained at a permissive temperature: *mCherry* (n = 24) or *Rbf* (n = 16) shRNA expression – permitted. **D.** (i) Constitutive suppression of neuronal RBF expression via *tub*-Gal80^ts^ significantly decreases peak locomotor activity in *Rbf^120a^*; *nsyb* > *Rbf* adult males. *Rbf^120a^*, *nsyb* > *Rbf*: n = 13; *Rbf^120a^*, *tub*-Gal80^ts^, *nsyb* > *Rbf*: n = 10. (ii) Peak activity in adult male flies that robust RBF expression solely permitted in adult-stage post-mitotic neurons is not significantly different from *Rbf^120a^* hypomorphs with constitutive post-mitotic neuronal expression of RBF. *Rbf^120a^*, *nsyb* > *Rbf*: n = 15; *Rbf^120a^*, *tub*-Gal80^ts^, *nsyb* > *Rbf*: n = 13. **E.** Model of dynamic changes in RBF function in developing and adult neurons. Error bars: SEM. * p< 0.05, ** p<0.005, *** p< 0.0005, ns – p> 0.05, Kruskal-Wallis test with Dunn’s post-hoc test (A), unpaired t-test with Welch’s correction (Ci, Cii, Di, Dii).

To more precisely interrogate whether Rbf LOF affects adult behaviour due to developmental perturbations or cell-autonomous activity in adult neurons, we performed complementary adult-stage neuron-specific knockdown and rescue experiments. To do so we utilised *tub*-Gal80^ts^, a globally expressed temperature-sensitive inhibitor of Gal4-mediated transgene expression^40^. In concert with the *nsyb*-Gal4 driver and *Rbf* shRNA or transgenes, this construct allowed us to examine the effects of both adult neuron-specific *Rbf* knockdown in an otherwise wild-type background (Figure 7B, C), and adult neuron-specific re-expression of Rbf in an *Rbf* hypomorph background (Figure 7B, D). We first found that, as expected, peak locomotion in wild-type flies did not significantly differ from controls when *Rbf* shRNA expression in post-mitotic neurons was constitutively repressed at 22°C by active Gal80^ts^ (Figure 7B, Ci). In contrast, inducing adult-stage neuronal knockdown of *Rbf* by shifting mature experimental flies to 29°C (Gal80 inactive, *Rbf* shRNA expressed) significantly reduced peak locomotion compared to control flies expressing an irrelevant shRNA (Figure 7B, Cii).

In the converse experiment, *Rbf* hypomorphs expressing transgenic Rbf in post-mitotic neurons (*Rbf*^120^, *nsyb > Rbf*) showed significantly higher peak locomotion at 22°C compared to flies of the same genotype but harbouring the repressive *tub*-Gal80^ts^ construct (Figure 7B, Di; Gal80 active, transgenic Rbf not expressed). However, when flies were raised at 22°C and then moved to a permissive temperature of 29°C at the adult stage (Figure 7B, Dii; Gal80 inactive, Rbf expressed), we observed no difference in peak activity between these two genotypes, suggesting that adult-specific restoration of Rbf expression in post-mitotic neurons was sufficient to rescue locomotor defects in *Rbf* hypomorphs. By extension, these findings suggest that defects in post-mitotic neuronal function may contribute to morbidities in RBL2 patients, particularly those associated with motor dysfunction.

## Discussion

The findings presented in this study shed light on a rare recessive neurodevelopmental disorder associated with mutations in the *RBL2*. Alongside the other RB family members (RB1 and RBL1), RBL2 acts as a crucial transcriptional regulator, particularly influencing neuronal differentiation and the survival of post-mitotic neurons. Unlike RB1 and RBL1, RBL2 is uniquely associated with a neurodevelopmental phenotype, however, only six individuals have been documented so far^12–14^. The limited number of patients, along with the isolated nature of the reports without comparative analysis, restricted the possibility of defining the phenotypic and genotypic spectrum of this disorder.

To address this gap, we have identified and clinically characterized a cohort of twenty-eight patients from eighteen families carrying homozygous or compound heterozygous predicted loss of function variants in *RBL2*. Within this cohort, we identified fourteen novel variants, bringing the total count of disease-associated pathogenic variants to nineteen. All variants are predicted to cause LOF, suggesting that the disease phenotype is primarily caused by total loss of protein function rather than alteration of defined active sites or protein interactions. However, it is essential to acknowledge potential limitations, such as the possibility of missing rare novel missense variants in specific domains that could be associated with milder forms of the disease.

The clinical presentation of affected individuals is characterized by a range of neurological and developmental abnormalities. Notably, global developmental delay, and intellectual disability were uniformly observed in the cohort. Interestingly, most of the patients lacked acquisition of key milestones, like walking and speech development, highlighting the disease severity from very early stages of the disorder. Postnatal microcephaly was observed in a considerable proportion of patients, while head circumference was normal in almost all the patients at birth, suggesting a progressive disorder. Remarkably, the vast majority of patients presented stereotypies, variably associated with autism spectrum disorder (ASD) and aggressive behaviour. Whilst the present cohort of patients did have facial dysmorphism, our analysis did not suggest a recognizable facial ‘gestalt’.

Common neuroimaging features included cerebral atrophy with an antero-posterior gradient variably associated with white matter volume loss and corpus callosum hypoplasia. In addition, cerebellar atrophy was noted in the majority of *RBL2* patients. Considering the presence of postnatal microcephaly in most cases, these findings suggest that neurodegeneration is an important feature of this disorder. As further confirmation, we also noted in most cases bilateral faint-to-marked signal changes at the level of the forceps minor, in keeping with an “ear-of-the-lynx” sign. This neuroimaging feature has been reported in hereditary spastic paraplegias (SPG7, 11 and 15)^41,42^ and other neurodegenerative disorders, including those related to variants in the *LNPK*, *CAPN1*, and *ATP13A2*^43–45^. Indeed, a neurodegenerative component is consistent with our *Drosophila* data, which suggest that the decreased brain size observed in *Rbf* hypomorphs is driven by an increase in cell death, most likely arising from cell-cycle defects in neuronal precursors and immature neurons.

Additionally, a total of three affected individuals were found to have expansile lesions: one orbital mass, one cystic mandibular lesion, and a large mass extending from the III ventricular floor to the prepontine cisterns. This is in line with previous studies that pointed to the potential role of RBL2 dysfunction in the evolution of cancer^46^ and confirms the dual role of RBL2 in both tumor suppression and neuronal differentiation and survival, thus providing further connection between tumorigenic processes and neurodevelopmental disorders^47^. Overall, both the clinical and neuroradiological findings underscored substantial intrafamilial and interfamilial variations in phenotypic expressions and severity, revealing considerable complexity within and between families.

To investigate the mechanisms linking RBL2 to neuronal development and function, we turned to the fruit fly, *Drosophila melanogaster*. Reductions in eye and optic lobe size, in *Drosophila Rbf* mutants are analogous to the microcephaly, brain atrophy and optic nerve hypoplasia seen in *RBL2* patients. Furthermore, the presence of apoptotic cells in the *Drosophila* developing brain is consistent with these phenotypes and points towards a model in which cell death during development leads to the atrophy observed in patient brains. The movement phenotypes we observe in mutant animals also seem to recapitulate aspects of the patient symptoms (e.g. gait abnormalities).

The similarities between *Drosophila* and patient phenotypes, coupled with the conserved sequence and molecular interactors of Rbf, support the use of *Drosophila Rbf* mutants as a model to understand the patho-mechanisms underlying *RBL2*-linked disorders and mammalian neural Rb function more broadly. Indeed, we uncovered an unexpected movement-promoting role for *Drosophila* Rbf in adult post-mitotic neurons. How Rbf influences gene expression in post-mitotic neurons is unclear. In some post mitotic cells, Rbf has been shown to modulate gene expression outside of its canonical function in repressing cell-cycle genes (for example in controlling muscle differentiation)^48^. Therefore, it is conceivable that Rbf also coordinates undefined gene expression programmes in mature neurons. Alternatively, Rbf neuronal-knockdown phenotypes may arise from the canonical action of Rbf on cell-cycle gene promoters – for example, in sustaining the epigenetic environment that maintains cell-cycle gene repression. Defining how Rbf and RBL2 influence gene regulatory networks in the mature fly and mammalian nervous system will thus be productive lines of future enquiry.

Our *Drosophila* studies advocate a model in which Rbf – and by extension RBL2 – acts sequentially in neural precursors and post-mitotic neurons to promote normal brain morphology and locomotor activity respectively (Figure 7E). Taken together, our work provides a clinical and experimental foundation to understand genotype-phenotype linkages in an *RBL2*-linked neurodevelopmental disorder and suggests that restoring RBL2 expression through gene therapy approaches may mitigate some of the multifaceted morbidities caused by *RBL2* pLOF. While our study illustrates parallels between human and fly Rb protein function in the nervous system, it is possible that some divergence in function has occurred between the two species. The human genome contains three Rb genes and eight genes encoding interacting E2F transcription factors^49^. Therefore, the greater complexity of the RB/E2F network in humans could result in altered biological outcomes. However, the similarities observed between RBL2 patient phenotypes and *Drosophila Rbf* mutant phenotypes indicates that RBL2/Rbf protein function in the CNS is likely to be conserved across metazoan species. Future studies in *Drosophila* may thus yield further insights into the mechanisms underlying *RBL2* pathology and inform therapeutic directions.

## Data Availability

All data produced in the present study are available upon request to the authors

## Acknowledgements

We would like to thank the patients and the families for their participation in this study. We would like to thank members of the Jepson and Houlden groups for their feedback and support on this project. We also thank Diego Sainz de La Maza and the Amoyel lab (University College London) for fly stocks and helpful advice.

## Ethical declarations

Individuals and/or their legal guardians recruited for this study gave informed consent for their participation. This study received approval from the Review Boards and Bioethics Committees at University College London Hospital (project 06/N076). Permission for inclusion of their anonymized medical data in this cohort, including photographs, was obtained using standard forms at each local site by the responsible referring physicians.

## Figure legends

**Supplementary Figure 1. A-B.** Single-cell RNAseq-derived t-distributed stochastic neighbor embedding (t-SNE) plots showing *Rbf* (A) and *Rbf2* (B) expression, alongside markers for glial cells (*repo*) and post-mitotic neurons (*nsyb*), in the adult fly brain. Gene expression across adult brain cells is illustrated using *SCope* [1]. **C.** Schematic showing protocol to assess climbing ability in adult flies. **D.** Number of flies (out of n = 10) passing a given threshold (see Materials and Methods) as a measure of climbing ability. Control adult males: n = 5 replicates, *Rbf^120a^* hemizygotes: n = 6 replicates. Error bars: SEM. ** p<0.005, unpaired t-test with Welch’s correction. **E-F.** *Rbf*-Gal4 driven nuclear mCherry expression in the *Drosophila* larval (E) and adult (F) brain. Adult neuronal nuclei are counterstained with an antibody against ELAV in (F). Scale bar = 100 µm.

**Supplementary Figure 2. A-B.** Expression in post-mitotic neurons of two distinct shRNAs (denoted as 330256 (A) and 65929 (B)) targeting *Rbf* mRNA reduces peak activity in adult Drosophila males. A: n = 20-37. B: n = 27-35. **C.** Number of flies (out of n = 10) passing a given threshold as a measure of climbing ability. n = 6-20 replicates. **D-E.** Crawling ability in control or *Rbf^120a^* hypomorph 3^rd^ instar larvae (D) or larvae subject to *Rbf* knockdown in post-mitotic neurons versus driver/transgene alone controls (E). D: n = 10 per genotype. E: n = 6-9. **F-G.** Mean optic lobe size (F) and number of apoptotic (DCP1-positive) cells in adult male brains subject to *Rbf* knockdown in post-mitotic neurons versus driver/transgene alone controls. F: n = 8-14. G: n = 5-7. Error bars: SEM. * p< 0.05, ** p<0.005, *** p< 0.0005, ns – p> 0.05, one-way ANOVA with Dunnett post-doc test (A, B, G), Kruskal-Wallis test with Dunn’s post-hoc test (C, E, F), or unpaired t-test with Welch’s correction (D).

**Supplementary Figure 3**. Knockdown of *Rbf* in glutamatergic neurons, reduces total activity during ZT0-24 in adult males. n = 11-41. Upper significance notation is relative to *Rbf* shRNA alone controls, lower significance notation is relative to Gal4 driver alone controls. Error bars: SEM. * p< 0.05, ** p<0.005, *** p< 0.0005, ns – p> 0.05, Kruskal-Wallis test with Dunn’s post-hoc test.

## Video legend

Video illustrating the stereotypies observed in patients with a neurodevelopmental encephalopathy due to biallelic *RBL2* pathogenic variants. Segment 1 (patient from family F3) shows orofacial stereotypies with teeth grinding associated with hand clasping stereotypies. Segment 2 (from family F11) shows multiple hands stereotypies comprising hand squeezing, hand mouthing, finger wiggling and finger tapping, and multiple orofacial stereotypies with a prominent involvement of the tongue.

