## Supplementary figures and images for "Clinical and neurogenetic characterisation of autosomal recessive RBL2-associated progressive neurodevelopmental disorder"

### Supplementary Figure 1

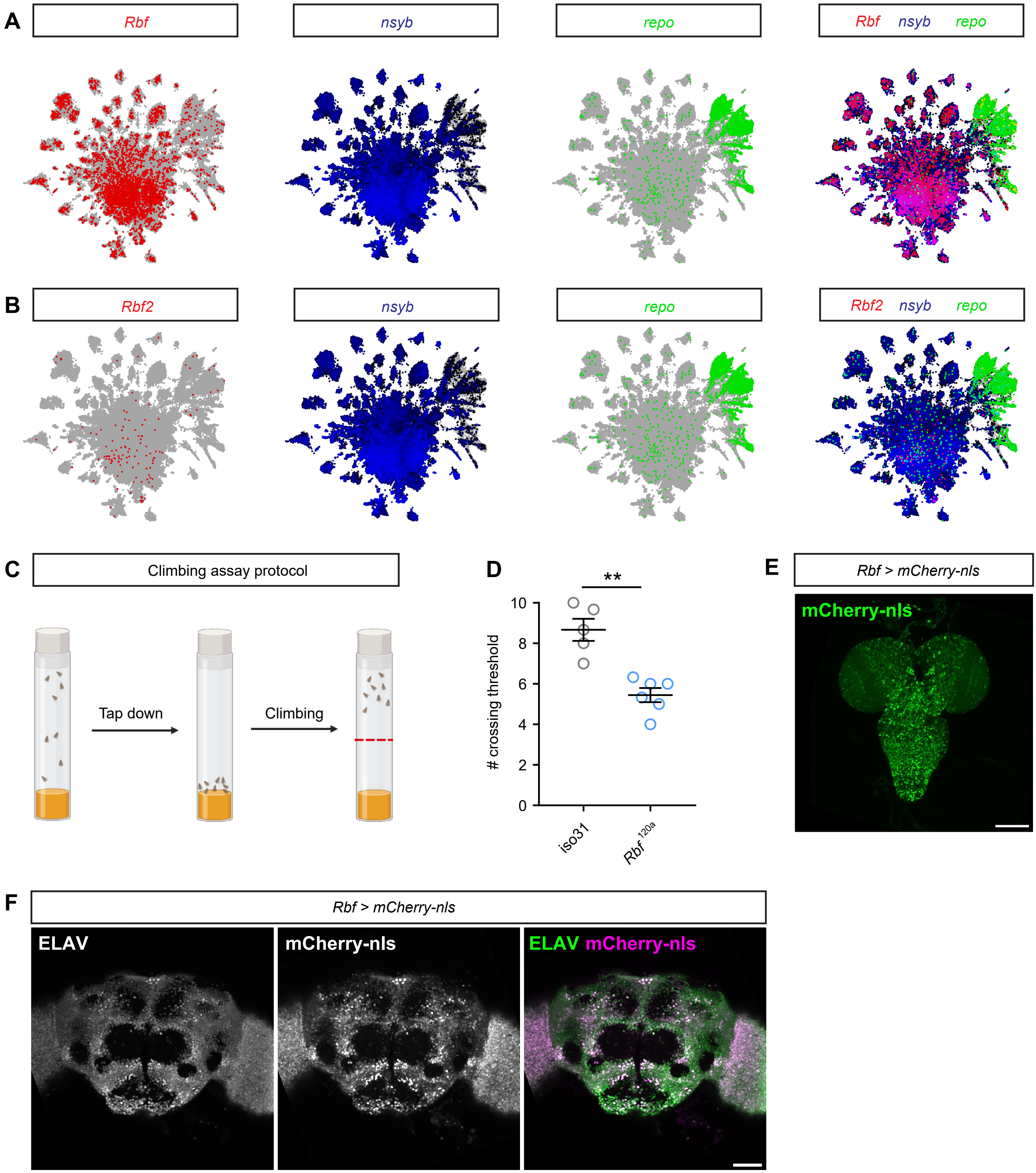

### Supplementary Figure 2

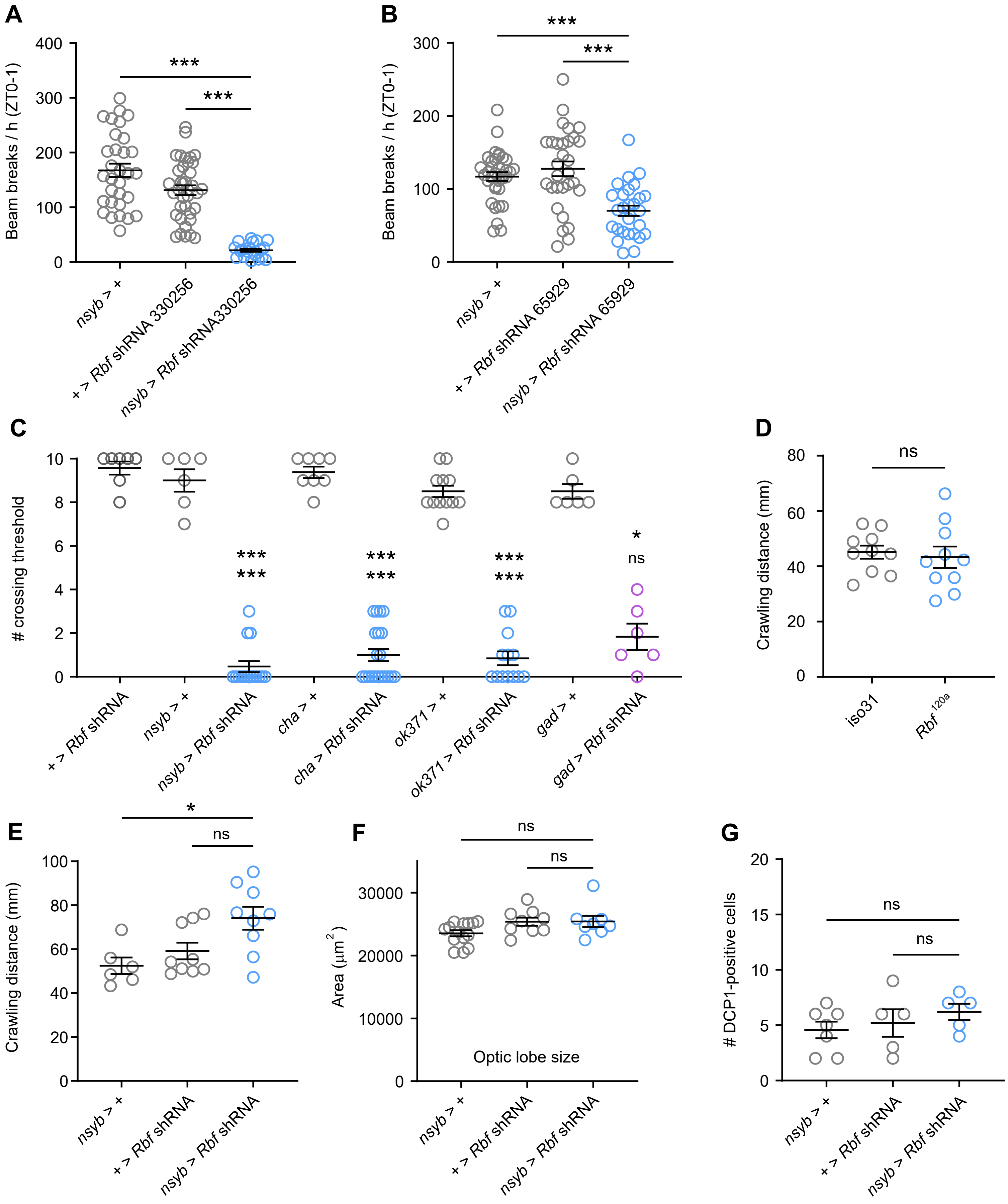

### Supplementary Figure 3

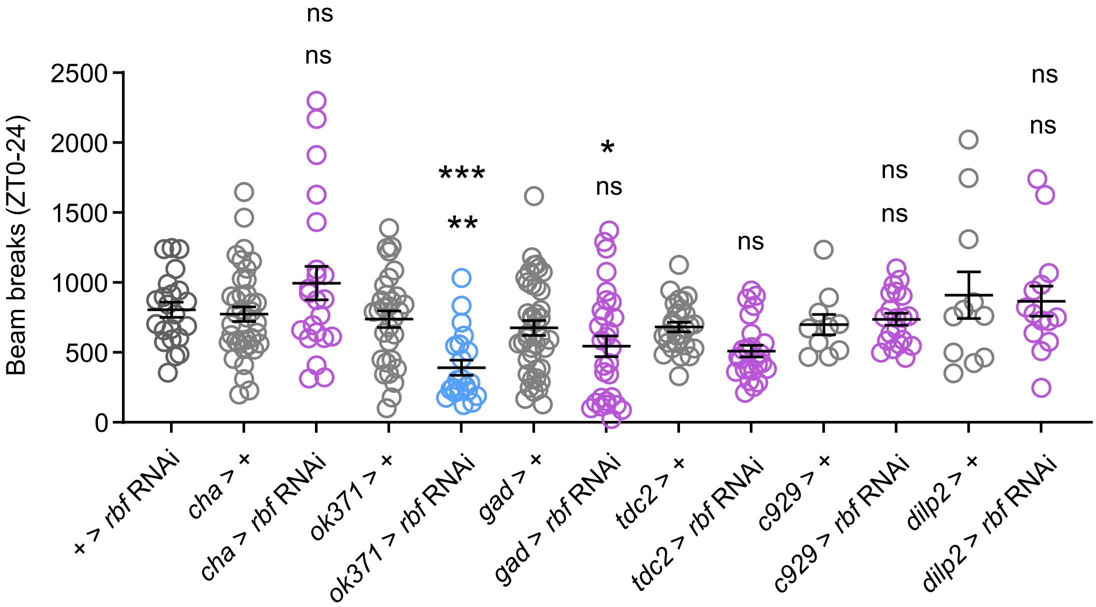
