## Supplementary Table 1 for "Clinical and neurogenetic characterisation of autosomal recessive RBL2-associated progressive neurodevelopmental disorder"

| **Genotype** | **Description** | **Source** |
| --- | --- | --- |
| *Rbf^120a^/FM7i, P{ActGFP}JMR3* | *Rbf* hypomorphic allele. | BDSC # 81612 |
| *Rbf^14^ w^1118^/FM7c* | *Rbf* null allele | BDSC # 7435 |
| *y^1^ sc^*^ v^1^ sev^21^; P{TRiP.HMS03004}attP2/TM3, Sb1* | RNAi targeting *Rbf.* | BDSC # 36744 |
| *P{VSH330256}attP40* | RNAi targeting *Rbf.* | VDRC # v330256 |
| *y^1^ sc^*^ v^1^ sev^21^; P{TRiP.HMC06195}attP2* | RNAi targeting *Rbf.* | BDSC # 65929 |
| *w^1118^_iso_*; *2_iso_*; *3_iso_* | Isogenised control strain (iso31) | Gift from Prof. Kyunghee Koh |
| *y^1^ TI{CRIMIC.TG4.2}^RbfCR00505-TG4.2^ w^*^/FM7h* | Expresses Gal4 under the control of *Rbf* regulatory sequences. | BDSC # 78934 |
| *w^1118^; P{GawB}VGlut^OK371^* | Expresses Gal4 in glutamatergic neurons. | Gift from Prof. Kyunghee Koh |
| *cha-Gal4* | Expresses Gal4 in cholinergic neurons. | Gift from Prof. Kyunghee Koh |
| *Gad-Gal4* | Expresses Gal4 in GABAergic neurons. | Gift from Prof. Kyunghee Koh |
| *Dilp2-Gal4* | Expresses GAL4 in insulinergic cells. | Gift from Prof. Kyunghee Koh |
| *C929-Gal4* | Expresses GAL4 in peptidergic cells. | Gift from Prof. Kyunghee Koh |
| *Tdc2-Gal4* | Expresses GAL4 in aminergic cells. | BDSC # 9313 |
| *w^*^; UAS-Rbf* | Expressed *Rbf* under the control of UAS sequence. | BDSC # 50746 |
| *Rbf^120a^/FM7i ; UAS-Rbf / CyO* | Allows for targeted expression of Rbf in *Rbf* hypomorph background. | This study |
| *tub-Gal80^ts^* | Ubiquitous temperature sensitive expression of the Gal80 repressor | Gift from Prof. Kyunghee Koh |
| *nSyb-Gal4* | Expresses Gal4 pan-neuronally. | BDSC#51635 |
| *tubGal80^ts^; nSyb-Gal4* | Allows for pan-neuronal temperature inducible expression. | This study |
| *UAS-mCherry-NLS* | Fluorescent reporter. | BDSC# 38424 |
| *y[1] sc[*] v[1] sev[21]; P{y[+t7.7] v[+t1.8]=UAS-mCherry.VALIUM10}attP2* | RNAi control. | BDSC# 35787 |
